# Alterations in Sarcopenia Index are Associated with Inflammation, Gut and Oral Microbiota among Heart Failure, Left Ventricular Assist Device and Heart Transplant Patients

**DOI:** 10.1101/2023.08.30.23294874

**Authors:** Melana Yuzefpolskaya, Bruno Bohn, Annamaria Ladanyi, Alberto Pinsino, Lorenzo Braghieri, Matthew R. Carey, Kevin Clerkin, Gabriel T. Sayer, Farhana Latif, Takeda Koji, Nir Uriel, Renu Nandakumar, Anne-Catrin Uhlemann, Paolo C. Colombo, Ryan T. Demmer

## Abstract

**Background:** Sarcopenia, characterized by loss of muscle mass and function, is prevalent in heart failure (HF) and associated with poor outcomes. We investigated alterations in sarcopenia index (SI), a surrogate marker of skeletal muscle mass, in HF, left ventricular assist device (LVAD) and heart transplant (HT) and assessed its relationship with inflammation and digestive tract (gut and oral) microbiota.

**Methods:** We enrolled 460 HF, LVAD and HT patients. Repeated measures pre and post procedures were obtained in a subset of LVAD and HT patients. Sarcopenia index (serum Creatinine/Cystatin C) and inflammatory biomarkers (C-reactive protein, interleukin-6, tumor necrosis factor-alpha) were measured in 271 and 622 blood samples, respectively. Gut and saliva microbiota were assessed via 16S rRNA sequencing among 335 stool and 341 saliva samples. Multivariable regression models were used to assess the relationship between SI and i) New York Heart Association class; ii) pre-vs. post-LVAD or HT; iii) biomarkers of inflammation and microbial diversity.

**Results:** Median (interquartile range) ln-SI was −0.13(−0.32,0.05). Ln-SI decreased across worsening HF class, further declined within the 1-month after LVAD and HT and rebounded over time to the levels of symptomatic HF. Ln-SI demonstrated an inverse correlation with inflammation (r=-0.28, p<0.0001), and a positive correlation with gut (r=0.26, p<0.0001) and oral microbial diversity (r=0.24, p<0.0001). Presence of the gut taxa *Roseburia inulinivorans* was associated with increased SI.

**Conclusions:** SI levels decreased in symptomatic HF and remained decreased long-term after LVAD and HT. SI levels covaried with inflammation, gut and oral microbiota in a similar fashion.

## INTRODUCTION

Heart Failure (HF) is a multi-faceted and life-threatening syndrome associated with significant mortality and morbidity^1^. Despite progress in therapeutics for chronic HF, disease remains progressive, and lifesaving treatment options for end-stage HF are limited to heart transplantation (HT) and left ventricular assist device (LVAD). HF progression is characterized by increase in congestion, inflammation, and oxidative stress, resulting in a clinical syndrome of end-stage HF commonly manifested as multiorgan system dysfunction and overall wasting continuum, including sarcopenia^2–4^. Sarcopenia is the skeletal muscle disorder characterized by a loss of muscle mass and function; it is strongly associated with HF prognosis^5^. The mechanisms promoting these inflammatory, oxidative responses and clinical sequalae including sarcopenia are incompletely understood. Additionally, sarcopenia diagnosis requires complex multimodal assessment of muscle quantity and quality, making clinical diagnosis challenging. Sarcopenia index (SI), based on serum creatinine (generated by skeletal muscle cells) and Cystatin C (produced by all nucleated cells), is a readily available biomarker that has been utilized as a reliable measure of skeletal muscle mass in several disease populations including critically ill^6^ and organ transplant recipients^7^.

The gastrointestinal tract contains a dynamic microbial community living in close contact with a substantial mucosal surface area. Recent evidence suggests gut and oral microbiome is an active contributor to HF pathogenesis and progression^8–10^. Specifically, imbalanced microbial communities, characterized by altered diversity (dysbiosis) with enrichment for pathogens and depletion of beneficial taxa (e.g., nitrate reducers and producers of short-chain fatty acids (SCFAs)), along with loss of intestinal mucosal barrier function, may worsen the disease by further inciting the inflammatory and oxidative milieu of HF. Several clinical studies including our own work, have reported associations between gut microbiome and HF phenotype^11–15^. More recently, animal models demonstrated a mechanistic link between gut microbial composition and skeletal muscle mass and function, commonly referred to as the *gut-muscle axis*^16, 17^. Limited human data exist highlighting reduced gut diversity and depletion of taxa with SCFA potential^18–20^ as well as taxa that contributes to the amino acid (AA) metabolism^21^ among sarcopenic patients, while no data is available among HF, LVAD and HT patients.

In a fashion not dissimilar to the gut microbiome, the oral microbiome has been repeatedly linked to a variety of systemic inflammatory diseases generally and to several cardiometabolic outcomes specifically^22, 23^. This concept has been most extensively studied in the context of periodontitis – a polymicrobial infection of the periodontal tissues, driven by alterations in the oral microbiome^24, 25^. While very few studies have examined the evidence linking periodontitis to HF risk, we have recently reported that periodontitis was strongly associated with incident HF^26^. Similarly, prior studies have reported an association between oral diseases and sarcopenia^27^.

Herein, in a large cohort of HF, LVAD and HT patients, we sought to i) describe variations in sarcopenia index across HF spectrum (New York Heart Association (NYHA) functional class (I-IV)), and at multiple time points after LVAD and HT; ii) describe gut and oral microbial changes across HF spectrum, and at multiple time points after LVAD and HT; and iii) investigate the relationship between SI and biomarkers of inflammation, gut and oral microbial diversity, with a particular focus on taxa that are proposed to participate in SCFA production and AA synthesis pathways.

## METHDOS

### Study Population

All patients were enrolled from June 2016 to February 2019 at Columbia University Irving Medical Center (CUIMC). Inclusion and exclusion criteria are provided in **Supplemental**. Patients were classified into the following categories: HF Class I, II, III, IV; post-LVAD: 1 month, 3 months, 6 months, 12 months; or post-HT: 1 week, 1 month, 3 months, 6 months, 12 months. Demographic and clinical information was extracted from electronic medical records (EMRs). Antibiotics use (peri-procedural and for treatment) 1 month before stool and/or blood sample collection was recorded from EMRs. The CUIMC Institutional Review Board approved the study protocol.

### Measurements of Serum Biomarkers

Biomarkers of inflammation (C-reactive protein [CRP], interleukin-6 [IL-6], tumor necrosis factor-a [TNF-a]) were measured in plasma as detailed in the **Supplemental**. Cystatin C was measured in serum using a particle enhanced immune turbidimetric assay (Cobas Integra 400 plus; Roche Diagnostics, Indianapolis, IN). Serum Creatinine was measured by enzymatic method (Cobas 8000; Roche Diagnostics, Indianapolis, IN). SI was calculated as serum creatinine value/Cystatin C value.

### Stool and Saliva analysis

Patients provided stool samples in sterile stool hats^15^. Details on stool and saliva collection and DNA extraction and 16S rRNA sequencing are provided in the **Supplemental**. A total of 394 of 595 stool samples had a corresponding blood sample collected within 30 days (median difference between blood and stool collection dates was 2 days). Saliva samples were collected during clinical visits or in an inpatient setting. A total of 435 of 522 saliva samples had a corresponding blood sample collected within 30 days (median difference between blood and saliva collection dates was 0 days).

### Chest Computed Tomography (CT) Analysis of Muscle Mass

Quantification of muscle mass was performed in a subgroup of patients with corresponding SI measures, using CT-measured pectoralis muscle area^28^. Further details regarding pectoralis muscle measurements are provided in **Supplemental.**

### Statistical Analysis

Demultiplexed sequence files were processed in R version 4.2.2, using the DADA2 pipeline to identify exact sequence variants (ESV)^29^. 16S analyses were carried out using the Phyloseq package. Alpha diversity (i.e., number and distribution of bacterial taxa within samples) was defined using the Shannon Index. Microbial diversity measures and log-transformed biomarkers of sarcopenia and inflammation were regressed on patient categories (NYHA Class, time post-LVAD or HT) separately by phenotype (HF, LVAD, or HT). The analyses utilized generalized linear mixed effects models with patients treated as random effects to account for repeated sampling of LVAD and HT patients. The same general regression approach was used in models assessing the relationship between gut microbiome or inflammation and SI. Adjusted and unadjusted least-square mean values of outcomes were computed across levels of any categorical variables. Models were adjusted for age, sex, race/ethnicity, and antibiotic use. Rare phyla (average relative abundance <0.5% were grouped as “Other”). DESeq2^30^ examined the association between patient category, SI, or inflammation and phylum level relative abundance (dependent variable). Analyses at the level of genus or species were conducted by operationalizing taxa level variables as presence vs. absence and then regressing SI values on this dichotomous variable. Taxa present in less than 10% or more than 90% of samples were excluded from analyses. A false discovery rate was used to correct for multiple comparisons. We present results for taxa that had an FDR<0.05 or those selected *a priori* based on a literature review of genera with known AA metabolism or SCFA synthesis functionality. The SI was computed as the natural log-transformed ratio of sCr to Cys-C. The *Inflammatory Score* was computed as the average between Z-score normalized, natural log-transformed CRP, TNF-⍺, and IL-6. Scores were computed for all patients with SI data and with data for at least two inflammatory biomarkers.

## RESULTS

### Baseline characteristics

We enrolled 460 HF, LVAD and HT patients. Baseline characteristics of patients providing blood samples are reported in **Table 1 through 3**, and characteristics of patients providing stool and saliva samples are provided in **Supplemental Table S1** and **S2**. Among 460 patients: i) 271 had Cys-C and sCr data available to calculate the SI (7 HF Class I, 20 Class II, 34 Class III, 85 Class IV, 90 LVAD and 127 HT); ii) 335 had gut microbiota data (8 HF Class I, 36 Class II, 45 Class III, 72 Class IV, 100 LVAD and 145 HT); and iii) 341 had saliva microbiota data (11 HF Class I, 42 Class II, 59 Class III, 55 Class IV, 111 LVAD and 124 HT).

**Table 1:**
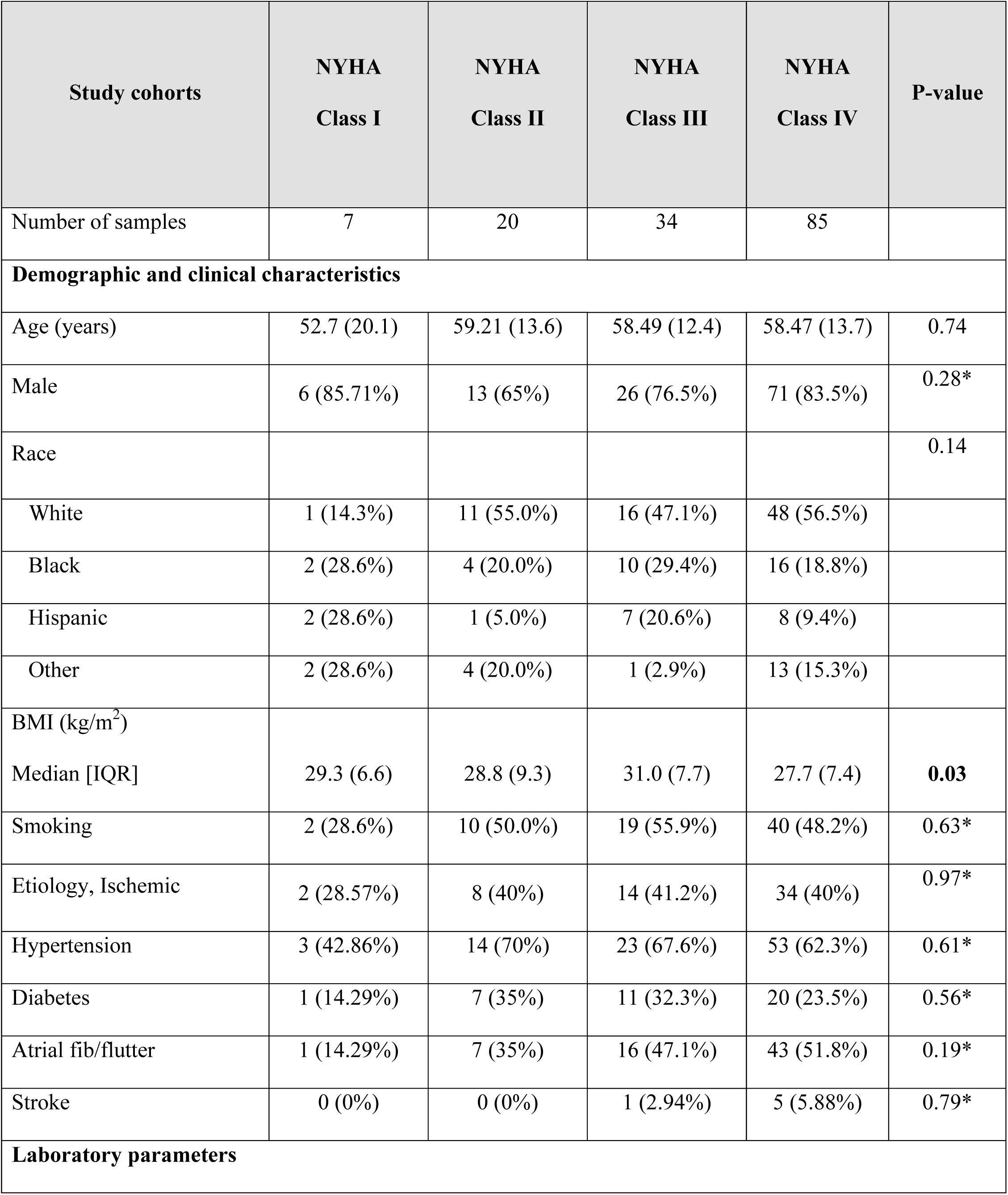
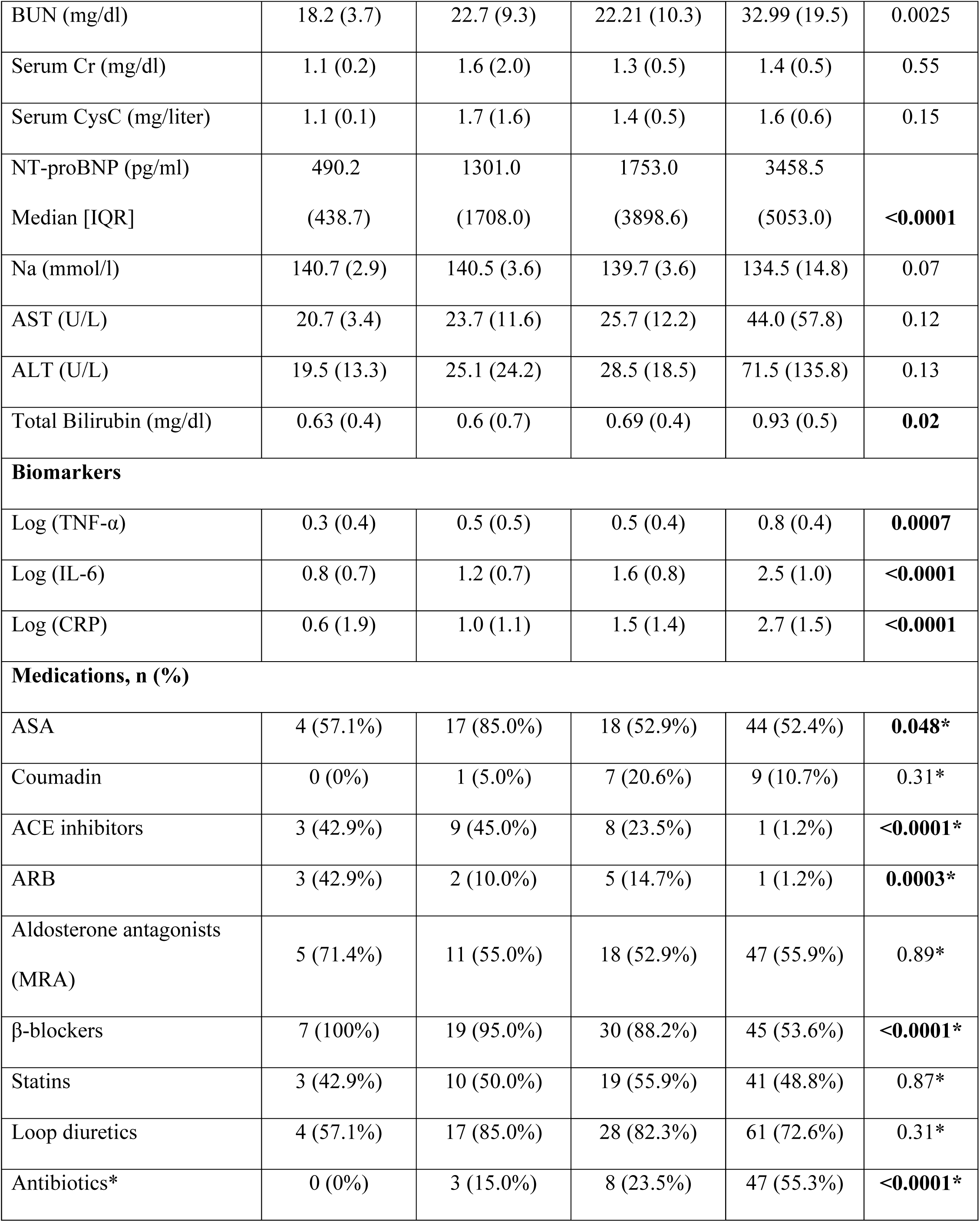
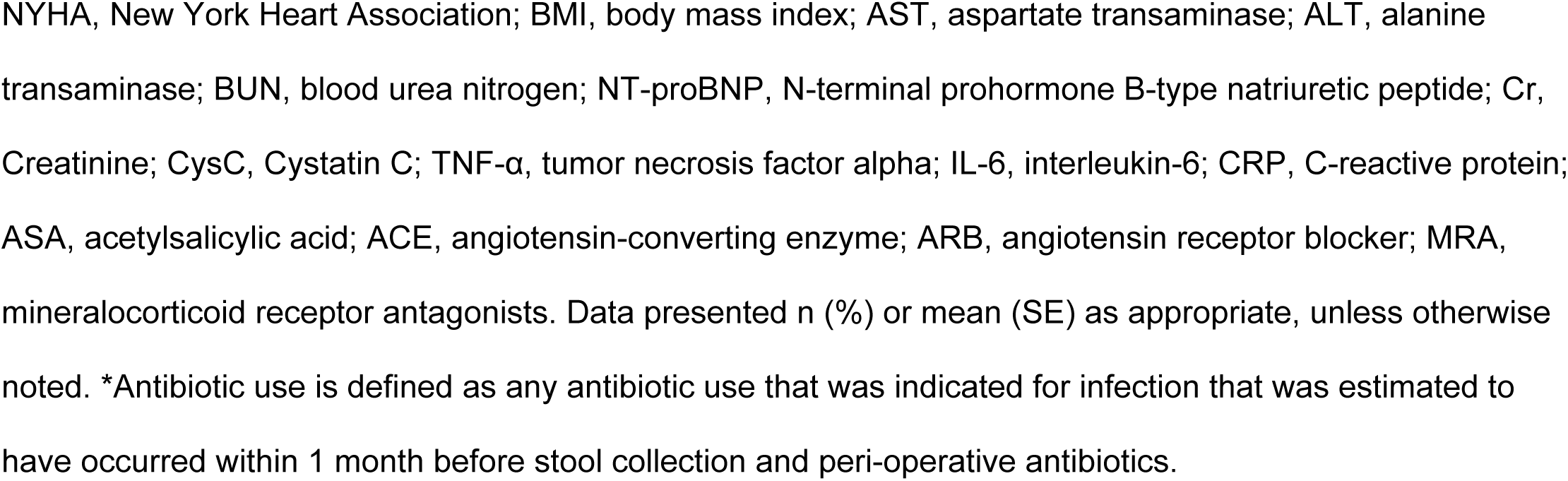
Baseline Characteristics of Heart Failure Patients Providing Blood Samples.

**Table 2:** Baseline Characteristics of LVAD Patients Providing Blood Samples.

| Study cohorts | LVAD<br>(1mo) | LVAD<br>(3mo) | LVAD<br>(6mo) | LVAD<br>(12mo) | LVAD<br>(>1yr) | P-value |
| --- | --- | --- | --- | --- | --- | --- |
| Number of samples | 46 | 15 | 27 | 19 | 22 |  |
| **Demographics and clinical characteristics** |  |  |  |  |  |  |
| Age (years) | 59.1 (12.7) | 63.6 (8.3) | 55.1 (14.6) | 54.6 (14.3) | 57.7 (15.0) | 0.25 |
| Male sex | 38 (82.6%) | 13 (86.7%) | 22 (81.5%) | 17 (89.5%) | 21 (95.4%) | 0.61* |
| Race |  |  |  |  |  | 0.53 |
| White | 29 (63.0%) | 10 (66.7%) | 15 (55.6%) | 11 (57.9%) | 12 (54.5%) |  |
| Black | 6 (13.0%) | 4 (26.7%) | 8 (29.6%) | 6 (31.6%) | 7 (31.8%) |  |
| Hispanic | 6 (13.0%) | 0 (0%) | 3 (11.1%) | 2 (10.5%) | 3 (13.6%) |  |
| Other | 5 (10.9%) | 1 (6.7%) | 1 (3.7%) | 0 (0%) | 0 (0%) |  |
| BMI (kg/m<sup>2</sup>) |  |  |  |  |  |  |
| Median [IQR] | 26.3 (5.7) | 25.5 (4.6) | 28.4 (6.8) | 27.9 (7.3) | 28.9 (6.5) | 0.21 |
| Smoking | 29 (63.0%) | 8 (53.3%) | 15 (55.6%) | 8 (42.1%) | 9 (40.9%) | 0.39* |
| Etiology, ischemic | 28 (60.9%) | 8 (53.3%) | 11 (40.7%) | 5 (26.3%) | 10 (45.4%) | 0.12* |
| INTERMACS Profile |  |  |  |  |  | 0.48 |
| 1 | 7 (15.2%) | 2 (13.3%) | 3 (11.5%) | 2 (10.5%) | 1 (4.8%) |  |
| 2 | 25 (54.3%) | 8 (53.3%) | 13 (50.0%) | 11 (57.9%) | 8 (38.1%) |  |
| 3 | 11 (23.9%) | 3 (20.0%) | 6 (23.1%) | 6 (31.6%) | 7 (33.3%) |  |
| 4 | 3 (6.5%) | 2 (13.3%) | 2 (7.7%) | 0 (0%) | 2 (9.5%) |  |
| 5-7 or not provided | 0 (0%) | 0 (0%) | 2 (7.7%) | 0 (0%) | 3 (14.3%) |  |
| Hypertension | 30 (65.2%) | 10 (66.7%) | 17 (63.0%) | 12 (63.2%) | 12 (54.5%) | 0.94* |
| Diabetes | 16 (34.8%) | 5 (33.3%) | 12 (44.4%) | 7 (36.8%) | 6 (27.3%) | 0.81* |
| Atrial fib/flutter | 19 (41.3%) | 5 (33.3%) | 13 (48.1%) | 9 (47.4%) | 10 (45.4%) | 0.90* |
| Stroke | 2 (4.3%) | 0 (0%) | 3 (11.1%) | 3 (16.7%) | 1 (4.5%) | 0.30* |
| Time after LVAD,<br>months | 20 | 103 | 188 | 340.0 | 1177.5 |  |
| Median [IQR] | (10.4) | (13) | (67.5) | (53.0) | (824.5) | **<0.0001** |
| **Laboratory parameters** |  |  |  |  |  |  |
| BUN (mg/dl) | 19.1 (13.0) | 28.53 (15.1) | 25.44 (17.3) | 24.2 (11.6) | 21.7 (10.0) | 0.13 |
| Serum Cr (mg/dl) | 1.2 (0.5) | 1.5 (0.6) | 1.3 (0.4) | 1.8 (1.4) | 1.4 (0.5) | **0.02** |
| Serum CysC (mg/l) | 1.6 (0.6) | 1.8 (0.8) | 1.5 (0.5) | 1.8 (1.1) | 1.3 (0.4) | 0.07 |
| NT-proBNP (pg/ml) | 3177.0 | 1915.0 | 1038.0 | 905.2 | 617.1 |  |
| Median [IQR] | (3071.5) | (1838.2) | (1529.2) | (1143.5) | (1289.7) | **<0.0001** |
| Na (mmol/l) | 136.2 (3.9) | 140.2 (3.3) | 139.8 (3.7) | 140.3 (2.4) | 140.6 (2.5) | **<0.0001** |
| AST (U/L) | 26.8 (12.0) | 28.3 (13.8) | 27.3 (22.2) | 22.3 (5.9) | 24.5 (6.1) | 0.65 |
| ALT (U/L) | 26.37 (17.0) | 22.7 (9.9) | 21.7 (12.1) | 18.3 (9.1) | 20.5 (7.9) | 0.17 |
| **Biomarkers** |  |  |  |  |  |  |
| Log (TNF-α) | 0.9 (0.4) | 0.9 (0.5) | 0.6 (0.3) | 0.5 (0.6) | 0.4 (0.4) | **0.0003** |
| Log (IL-6) | 3.0 (0.7) | 1.8 (0.8) | 1.7 (0.7) | 1.7 (0.6) | 1.3 (1.0) | **<0.0001** |
| Log (CRP) | 3.9 (0.8) | 1.91 (1.3) | 1.8 (1.0) | 1.6 (1.2) | 1.8 (1.1) | **<0.0001** |
| **Medications, n (%)** |  |  |  |  |  |  |
| ASA | 39 (86.7%) | 11 (73.3%) | 18 (66.7%) | 13 (68.4%) | 16 (72.7%) | 0.25* |
| Coumadin | 29 (64.4%) | 14 (93.3%) | 25 (92.6%) | 17 (89.5%) | 17 (77.3%) | **0.02\*** |
| ACE inhibitors | 5 (10.9%) | 4 (26.7%) | 12 (44.4%) | 5 (26.3%) | 5 (22.7%) | **0.03\*** |
| ARB | 0 (0%) | 1 (6.7%) | 4 (14.8%) | 4 (21.0%) | 3 (13.6%) | **0.01\*** |
| Aldosterone<br>Antagonists<br>(MRA) | 19 (41.3%) | 6 (40.0%) | 15 (55.6%) | 5 (26.3%) | 7 (31.8%) | 0.32* |
| β-blockers | 22<br>(47.83%) | 13<br>(86.67%) | 22<br>(81.48%) | 19<br>(100%) | 19<br>(86.36%) | **<0.001\*** |
| Statins | 12 (26.09%) | 3 (20%) | 7 (25.93%) | 2 (10.53%) | 5 (22.73%) | 0.7275* |
| Loop Diuretics | 27<br>(60%) | 11<br>(73.33%) | 21<br>(77.78%) | 13<br>(68.42%) | 12<br>(54.55%) | 0.4127* |
| *Antibiotics | 45 (97.83%) | 2 (13.33%) | 10 (37.04%) | 4 (21.05%) | 11 (50%) | **<0.0001\*** |

LVAD, Left Ventricular Assist Device; BMI, body mass index; AST, aspartate transaminase; ALT, alanine transaminase; BUN, blood urea nitrogen; NT-proBNP, N-terminal prohormone B-type natriuretic peptide; Cr, Creatinine; CysC, Cystatin C; TNF-α, tumor necrosis factor alpha; IL-6, interleukin-6; CRP, C-reactive protein; ASA, acetylsalicylic acid; ACE, angiotensin-converting enzyme; ARB, angiotensin receptor blocker; MRA, mineralocorticoid receptor antagonists. Data presented n (%) or mean (SE) as appropriate, unless otherwise noted. \*Antibiotic use is defined as any antibiotic use that was indicated for infection that was estimated to have occurred 1 month before stool collection and peri-operative antibiotics.

**Table 3:** Baseline Characteristics of HT Patients Providing Blood Samples.

| Study cohorts | HT<br>(1wk) | HT<br>(1mo) | HT<br>(3mo) | HT<br>(6mo) | HT<br>(12mo) | HT<br>(>1yr) | P-value |
| --- | --- | --- | --- | --- | --- | --- | --- |
| Number of samples | 62 | 77 | 75 | 60 | 37 | 36 |  |
| **Demographic and clinical characteristics** |  |  |  |  |  |  |  |
| Age, (years) | 54<br>(12.53) | 53.15<br>(12.7) | 53.78<br>(12.8) | 53.52<br>(13.06) | 53.5<br>(11.41) | 59.41<br>(10.48) | 0.2054 |
| Male sex | 47<br>(75.81%) | 58<br>(75.32%) | 59<br>(78.67%) | 47<br>(78.33%) | 30<br>(81.08%) | 27<br>(75%) | 0.9802* |
| Race |  |  |  |  |  |  | 0.9441 |
| White | 35<br>(56.45%) | 41<br>(53.25%) | 39<br>(52%) | 30<br>(50%) | 21<br>(56.76%) | 12<br>(33.33%) |  |
| Black | 15<br>(24.19%) | 22<br>(28.57%) | 21<br>(28%) | 20<br>(33.33%) | 11<br>(29.73%) | 15<br>(41.67%) |  |
| Hispanic | 9<br>(14.52%) | 10<br>(12.99%) | 11<br>(14.67%) | 8<br>(13.33%) | 4<br>(10.81%) | 7<br>(19.44%) |  |
| Other | 3<br>(4.84%) | 4<br>(5.19%) | 4<br>(5.33%) | 2<br>(3.33%) | 1<br>(2.7%) | 2<br>(5.56%) |  |
| BMI, (kg/m2)<br>Median [IQR] | 26.91<br>(6.68) | 25.73<br>(5.42) | 26.15<br>(6.27) | 26.68<br>(7.39) | 27.76<br>(7.98) | 30.05<br>(10.55) | **0.0003** |
| Smoking | 27<br>(44.26%) | 31<br>(40.79%) | 36<br>(48.65%) | 25<br>(42.37%) | 18<br>(48.65%) | 15<br>(41.67%) | 0.9235* |
| Etiology, ischemic | 18<br>(29.03%) | 16<br>(20.78%) | 22<br>(29.33%) | 20<br>(33.33%) | 12<br>(32.43%) | 14<br>(38.89%) | 0.4018* |
| Hypertension | 31<br>(50%) | 43<br>(55.84%) | 46<br>(61.33%) | 36<br>(60%) | 23<br>(62.16%) | 34<br>(94.44%) | **0.0008** |
| Diabetes | 15<br>(24.19%) | 19<br>(24.68%) | 22<br>(29.33%) | 15<br>(25%) | 11<br>(29.73%) | 21<br>(58.33%) | **0.0053** |
| Atrial fib/flutter | 31<br>(52.54%) | 38<br>(51.35%) | 39<br>(52.7%) | 30<br>(50%) | 17<br>(45.95%) | 9<br>(25%) | 0.0993 |
| Stroke | 7<br>(11.67%) | 10<br>(13.33%) | 9<br>(12.16%) | 6<br>(10%) | 5<br>(13.51%) | 3<br>(8.33%) | 0.9756* |
| Time after HT,<br>months<br>Median [IQR] | 8<br>[2.72] | 22<br>[9.04] | 67.04<br>[30.9] | 154.04<br>[30.7] | 306<br>[47] | 1737.02<br>[2246.94] | **<0.0001** |
| **Laboratory parameters** |  |  |  |  |  |  |  |
| BUN (mg/dl) | 36.07<br>[19.5] | 30.88<br>[13.1] | 29.16<br>[12.2] | 28.36<br>[11.4] | 23.89<br>[9.98] | 26.34<br>[10.24] | **0.0004** |
| Serum Cr (mg/dl) | 1.25<br>[0.51] | 1.29<br>[0.91] | 1.34<br>[0.72] | 1.59<br>[1.19] | 1.37<br>[0.45] | 1.55<br>[1.17] | 0.1977 |
| Serum CysC (mg/l) | 1.89<br>[0.73] | 1.6<br>[0.85] | 1.49<br>[0.58] | 1.5<br>[0.74] | 1.32<br>[0.38] | 1.57<br>[0.95] | **0.0032** |
| Na (mmol/l) | 138.39<br>[4.44] | 138.58<br>[2.78] | 140.42<br>[3.62] | 141.73<br>[2.64] | 141.71<br>[1.96] | 142.49<br>[2.63] | **<0.0001** |
| AST (U/L) | 23.61<br>[9.71] | 22.09<br>[11.47] | 25.59<br>[16] | 25.1<br>[9.38] | 25.91<br>[8.92] | 22.75<br>[9.05] | 0.3688 |
| ALT (U/L) | 31.72<br>[22.56] | 33.8<br>[31.96] | 27.2<br>[21.51] | 24.73<br>[21.38] | 22.69<br>[16.82] | 22.39<br>[12.5] | **0.0468** |
| Total Bilirubin<br>(mg/dl) | 0.76 | 0.56 | 0.46 | 0.55 | 0.54 | 0.61 | **0.0006** |
|  | [0.47] | [0.43] | [0.24] | [0.36] | [0.23] | [0.44] |  |
| **Biomarkers** |  |  |  |  |  |  |  |
| Log (TNF- $\alpha$) | 0.22<br>(0.52) | 0.09<br>(0.47) | 0.28<br>(0.49) | 0.48<br>(0.46) | 0.36<br>(0.53) | 0.51<br>(0.35) | **0.0014** |
| Log (IL-6) | 2.49<br>(1.01) | 1.6<br>(0.88) | 1.32<br>(1.05) | 1.41<br>(1.1) | 1.21<br>(0.96) | 1.39<br>(0.61) | **<0.0001** |
| Log (CRP) | 2.78<br>(1.32) | 1.22<br>(1.59) | 0.5<br>(1.54) | 0.89<br>(1.65) | 0.97<br>(1.28) | 1.25<br>(1.05) | **<0.0001** |
| **Medications, n (%)** |  |  |  |  |  |  |  |
| ASA | 19<br>(30.65%) | 56<br>(72.73%) | 62<br>(82.67%) | 54<br>(90%) | 32<br>(86.49%) | 34<br>(94.44%) | **<0.0001** |
| Coumadin | 0 (0%) | 0 (0%) | 0 (0%) | 0 (0%) | 1 (2.7%) | 1 (2.8%) | **0.04\*** |
| ACE inhibitors | 0<br>(0%) | 1<br>(1.3%) | 0<br>(0%) | 2<br>(3.33%) | 1<br>(2.7%) | 2<br>(5.56%) | 0.13* |
| ARB | 0<br>(0%) | 0<br>(0%) | 2<br>(2.67%) | 1<br>(1.67%) | 3<br>(8.11%) | 13<br>(36.11%) | **<0.0001\*** |
| Aldosterone<br>Antagonists<br>(MRA) | 0 (0%) | 1 (1.3%) | 0 (0%) | 0 (0%) | 1 (2.7%) | 1 (2.8%) | 0.20* |
| β-blockers | 0<br>(0%) | 8<br>(10.39%) | 8<br>(10.67%) | 7<br>(11.67%) | 4<br>(10.81%) | 10<br>(27.78%) | **0.0023** |
| Statins | 27<br>(43.55%) | 67<br>(87.01%) | 69<br>(92%) | 54<br>(90%) | 33<br>(89.19%) | 31<br>(86.11%) | **<0.0001** |
| Loop diuretics | 24<br>(38.71%) | 21<br>(27.27%) | 27<br>(36%) | 15<br>(25%) | 3<br>(8.11%) | 10<br>(27.78%) | **0.0223** |
| Antibiotics* | 62<br>(100%) | 76<br>(98.7%) | 48<br>(64%) | 28<br>(46.67%) | 17<br>(45.95%) | 8<br>(22.22%) | **<0.0001** |
| **Immunosuppression, n (%)** |  |  |  |  |  |  |  |
| Tacrolimus | 59<br>(95.2%) | 75<br>(97.4%) | 74<br>(98.7%) | 58<br>(96.7%) | 34<br>(91.8%) | 25<br>(69.4%) | **<0.0001\*** |
| Prednisone | 57 (91.9%) | 75 (97.4%) | 74 (98.7%) | 59 (98.3%) | 34 (91.9%) | 24 (66.7%) | **<0.0001** |
| Mycophenolic Acid | 1 (1.6%) | 3 (3.9%) | 5 (6.7%) | 5 (8.3%) | 4 (10.8%) | 5 (13.9%) | 0.1315* |
| Mycophenolate<br>Mofetil | 60 (96.8%) | 70 (90.9%) | 64 (85.3%) | 44 (73.3%) | 26 (70.3%) | 23 (63.9%) | **<0.0001** |

HT, heart transplant; BMI, body mass index; AST, aspartate transaminase; ALT, alanine transaminase; BUN, blood urea nitrogen; NT-proBNP, N-terminal prohormone B-type natriuretic peptide; Cr, Creatinine; CysC, Cystatin C; TNF- $\alpha$ , tumor necrosis factor alpha; IL-6, interleukin-6; CRP, C-reactive protein; ASA, acetylsalicylic acid; ACE, angiotensin-converting enzyme; ARB, angiotensin receptor blocker; MRA, mineralocorticoid receptor antagonist. Data presented n (%) or mean (SE) as appropriate, unless otherwise noted. \*Antibiotic use is defined as any antibiotic use that was indicated for infection or prophylaxis as per HT protocol that was estimated to have occurred 1 month before stool collection and perioperative antibiotics.

All patients studied were predominantly white male, with no difference in age or HF etiology across HF categories. Of note, the proportion of LVAD patients treated with statins was significantly lower than HT or HF.

### Association of Sarcopenia Index with CT-measured Pectoralis Muscle Index (PMI)

In a subgroup of 38 HF, LVAD and HT patients with SI and chest CT data available within 0-14 days of SI measurement (median of 3 days), a correlation analysis between PMI (cm^2^/m^2^) and ln-SI was performed (r=0.46 (0.25-0.62), p<0.0001) (**Figure S1**).

### Variation in Sarcopenia Index Across Patient Categories

The median (interquartile range) ln-SI of the entire cohort was −0.13 (−0.32, 0.05) and ln-SI distributions across patient categories are shown in **Figure 1A**. In adjusted regression models (**Figure 1B**), mean ln-SI values decreased across worsening HF NYHA class; mean ln-SI values among HF class IV patients were significantly higher compared to LVAD 1 month (P<0.0001) and HT 1 week (P<0.0001) and HT 1 month (P<0.0001). Mean ln-SI values increased with time post-LVAD and HT but did not significantly differ from the levels observed among symptomatic HF patients.

**Figure 1:**
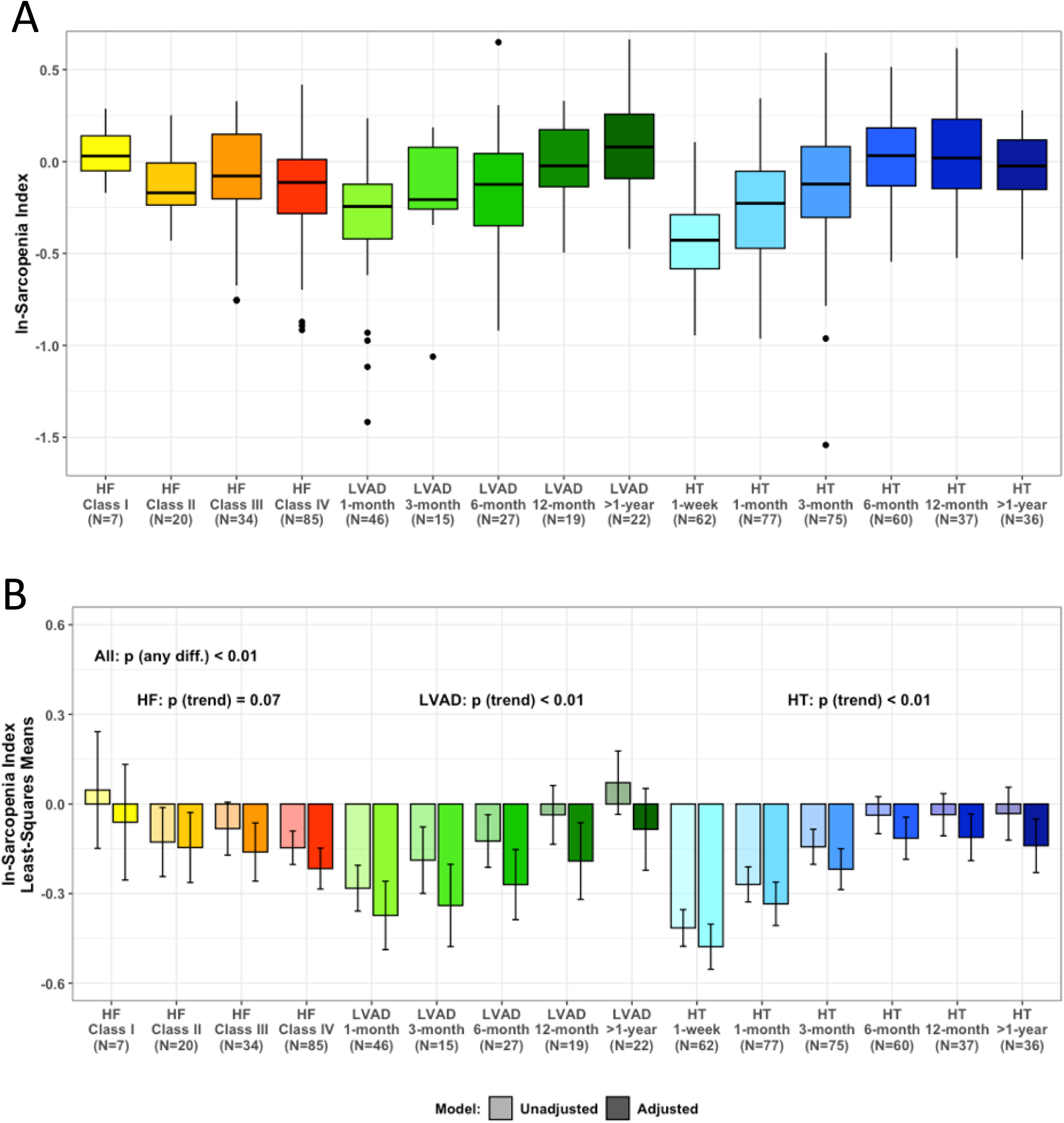
Variations in sarcopenia index (SI) across disease categories of heart failure (HF), left ventricular assist device (LVAD) and heart transplant (HT) patients. **A.** Unadjusted ln-transformed SI values (n=271). **B.** Unadjusted and adjusted least-squared means of ln-transformed SI values. Adjusted for age, sex, race/ethnicity, and antibiotics use. P-values correspond to adjusted models. NYHA indicates New York Heart Association.

### Variation in Gut and Oral Microbiota Across Patient Categories

Mean (SD) of the gut Shannon Index values for the entire cohort was 5.81±0.63. Variations in Shannon Index across patient category are shown in **Figure 2A**. After multivariable adjustment (**Figure 2B**), mean levels of the gut Shannon Index varied across all patient categories (p for any difference<0.0001), with a non-statistically significant trend for decreasing diversity across worsening HF NYHA class (p for trend=0.29). The lowest mean Shannon Index values were observed in the first month post-LVAD or HT, with a significant trend of increasing diversity with time after HT (p for trend<0.0001), but non-significant after LVAD (p for trend=0.46).

**Figure 2:**
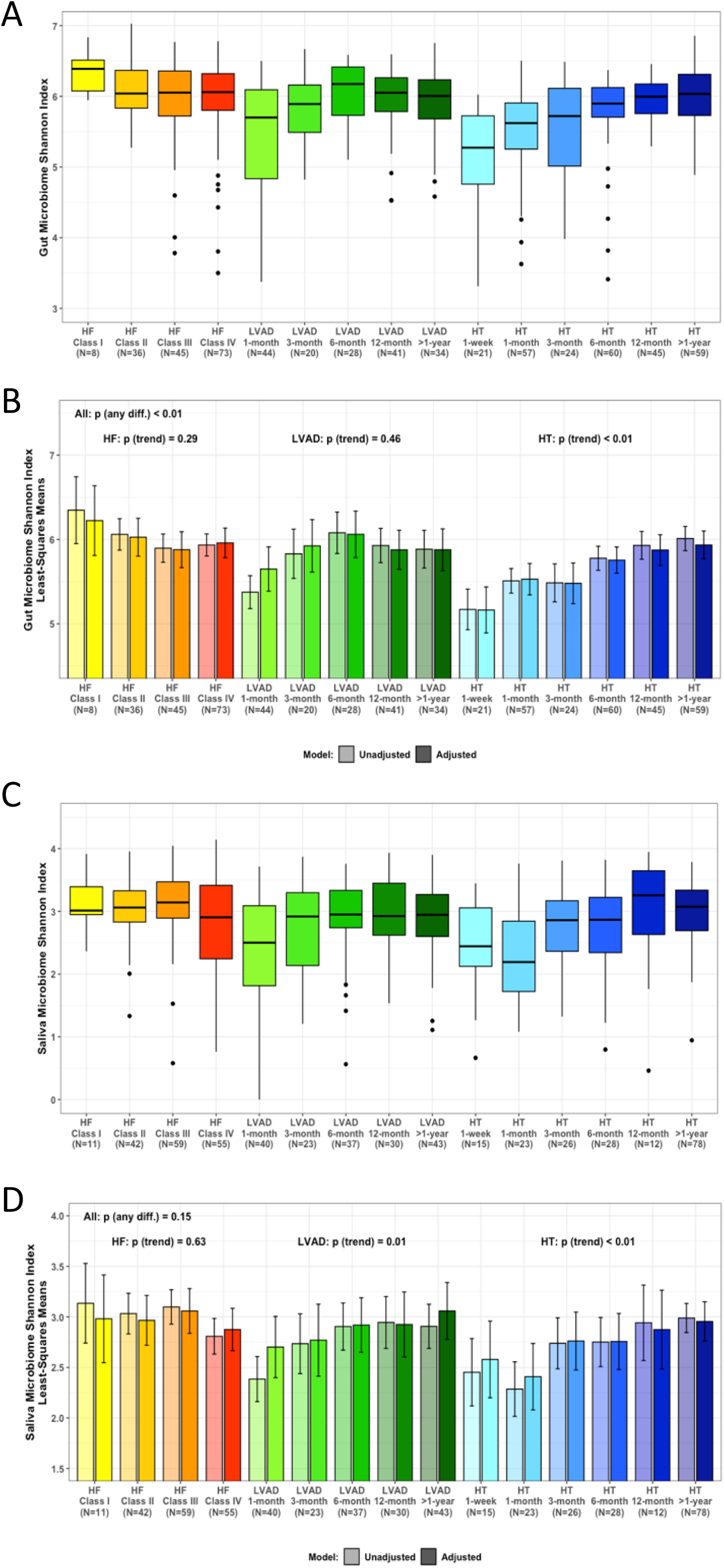
Measures of gut and saliva alpha diversity across patients’ categories. **A.** Unadjusted gut Shannon Index (n=335). **B.** Least-squared means of gut Shannon Index, adjusted for age, sex, race/ethnicity, and antibiotics use 1 month before stool collection. **C.** Unadjusted saliva Shannon Index (n=341). **D.** Least-squared means of saliva Shannon Index, adjusted for age, sex, race/ethnicity, and antibiotics use 1 month before stool collection. P-values correspond to adjusted models. NYHA, New York Heart Association; LVAD, left ventricular assist device; HT, heart transplant.

Mean value (SD) of the saliva Shannon Index was 2.84±0.70. Variations in the saliva Shannon Index across patient category are shown in **Figure 2C**. After multivariable adjustment (**Figure 2D**), no significant differences in mean levels of the Shannon Index were observed across all patient categories or across HF class. The lowest mean Shannon Index values were observed in the first month post-LVAD or HT, with a significant trend of increasing diversity with time after LVAD and HT.

In phylum level analyses of the gut microbiota average relative abundance appeared to increase for *Proteobacteria* and decrease for *Firmicutes* immediately following LVAD and HT with a temporal shift back towards the composition observed among HF patients (**Figure 3A**). In saliva phylum level analyses (**Figure 3B**), the phylum level relative abundance was less variable and did not appear to change dramatically even immediately following LVAD or HT. Overall, given the large variation in sample size across groups and the notable multiple comparisons issues, the power for statistical hypothesis testing is low and the observed patterns should be interpreted cautiously.

**Figure 3:**
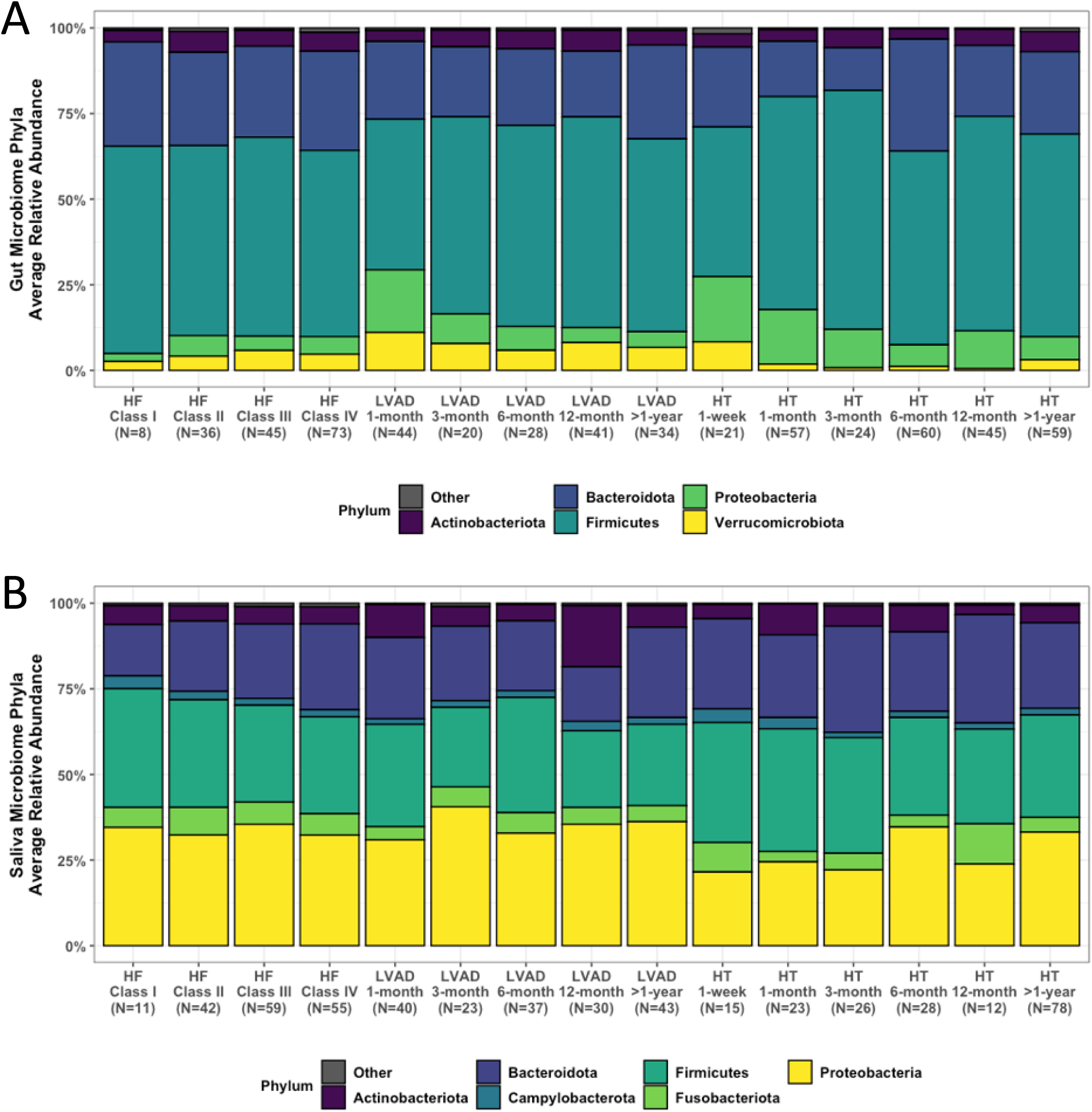
Average gut and saliva phyla relative abundance across patients’ categories. **A.** Gut phyla relative abundance (n=335). **B.** Saliva phyla relative abundance (n=341). Phyla with average relative abundance below 0.5% grouped as “Other”. NYHA, New York Heart Association; LVAD, left ventricular assist device; HT, heart transplant.

### Variation in Inflammatory Score Across Patient Category

Mean Inflammatory Score levels significantly differed between patient category (p for any difference < 0.0001, **Figure S2**). Increased inflammation was observed across worsening HF NYHA class (p for trend<0.0001), while inflammatory score declined with time post-LVAD (p for trend<0.0001) and HT (p for trend=0.005). Notably, the highest observed inflammatory score was early after LVAD. Individual variations in CRP, IL-6 and TNF-alpha are shown in **Figure S3.**

### Association Between Gut and Oral Shannon Index and Inflammatory Score

Associations between microbial diversity (oral and gut) and inflammation are presented in **Table S3**. Increased gut diversity is associated with lower inflammatory score while oral diversity was not.

### Association Between Sarcopenia Index, Inflammatory Score, Gut and Oral Shannon Index

The association between *ln-SI and the inflammatory score* among n=262 participants is shown in **Figure 4A**. When analyzing the entire cohort, ln-SI demonstrated a moderate inverse correlation with the inflammatory score (r = −0.28, p<0.0001). This association was observed within each subgroup. After multivariable adjustment in linear regression models, the inverse association between ln-SI and inflammatory score remained significant for the combined cohort and the HT subgroup, but not among HF or LVAD patients.

**Figure 4:**
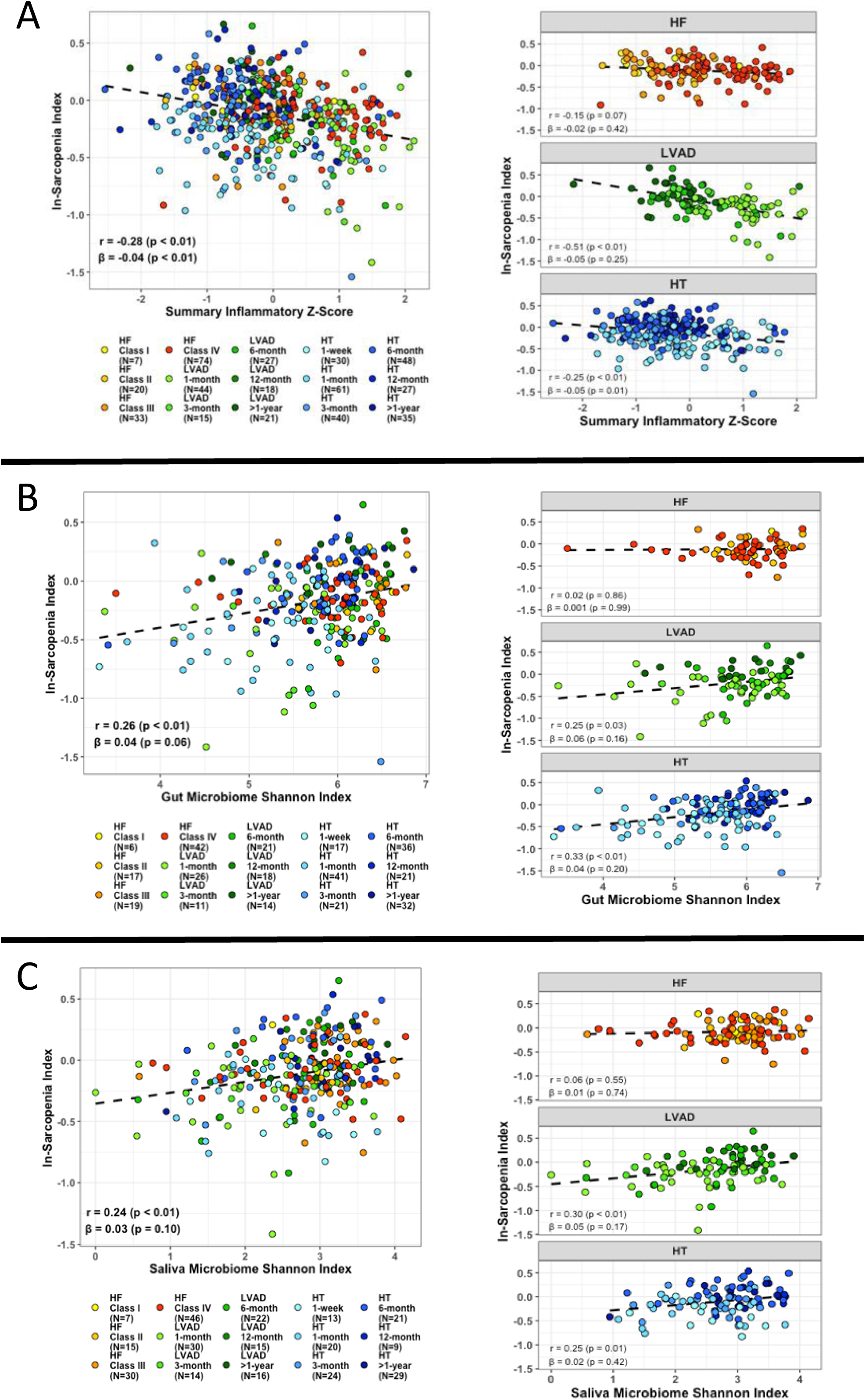
Association between sarcopenia index (SI) and inflammatory z-score, gut, and saliva alpha diversity for the entire cohort and for disease categories of heart failure (HF), left ventricular assist device (LVAD) and heart transplant (HT). **A.** Association between SI and inflammatory z-core for the entire cohort (n=262) and for individual cohorts of HF, LVAD and HT. **B.** Association between SI and gut Shannon Index for the entire cohort (N=170) and for individual cohorts of HF, LVAD and HT. **C.** Association between SI and saliva Shannon Index for the entire cohort (N=172) and for individual cohorts of HF, LVAD and HT. Adjusted models include patient category, age, sex, race/ethnicity, and antibiotic use. Unadjusted results present correlation coefficients and adjusted results present beta-coefficients. P-values correspond to adjusted models.

The association between *ln-SI and gut microbial diversity* among n=170 participants is shown in **Figure 4B**. A moderate positive correlation was observed in the entire cohort (r = 0.26, p<0.0001) and among LVAD and HT, but not among HF. This associations were attenuated and lost significance after multivariable adjustment in the combined and individual cohorts.

The association between *ln-SI and saliva microbial diversity* among n=172 participants is shown in **Figure 4C**. A moderate positive correlation was observed in the entire cohort (r = 0.24, p < 0.0001) and among LVAD and HT subgroups, but not among HF patients. These associations were not maintained after multivariable adjustment.

### Association between Sarcopenia Index and Gut and Oral Taxa

The top gut and oral taxa associated with SI are shown in **Figures 5A, B**, respectively. The presence (vs. absence) of the gut taxa *Roseburia inulinivorans* was associated with increased levels of SI after multivariable adjustment and these findings were also present in the subgroups of HF and HT patients. Additionally, there was a trend (not statistically significant) for lower SI levels among patients with *Eggerthella lenta* and *Klebsiella spp*. detected. Presence (vs. absence) of the oral taxa *Prevotella nanceiensis* and *Prevotella_7 spp*. was associated with higher levels of SI while presence of *Lacticaseibacillus spp*. and *Actinomyces naeslundii* was associated with lower levels of SI. Findings in patient subgroups are presented in **Figures S4 and S5.** Analyses focused on *a priori* identified taxa with potential AA synthesis functionality did not yield any notable findings (data not shown).

**Figure 5:**
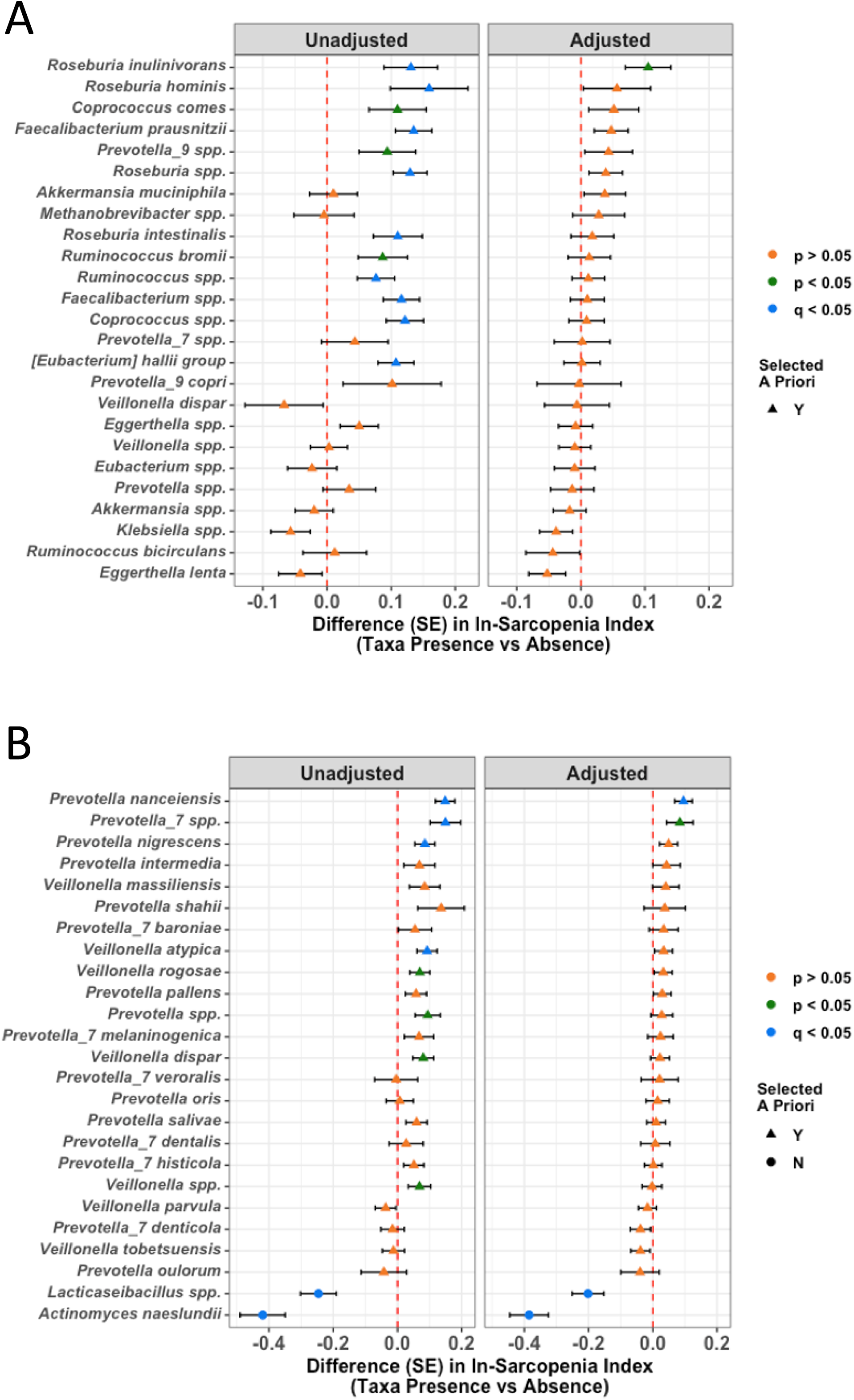
Change in sarcopenia index (SI) by presence vs absence of gut and saliva taxa among all patients. Taxa selected a priori to reflect genera known to produce/reduce amino acids and short chain fatty acids. Taxa present in less than 10% or more than 90% of samples were excluded. Adjusted models include patient category, age, sex, race/ethnicity, and antibiotic use. **A.** Change in SI and gut taxa. **B.** Change in SI and saliva taxa.

## DISCUSSION

Heart failure progression is associated with worsened SI, increased levels of inflammation and changes in gut and oral microbiota. Extreme digestive tract (gut and oral) dysbiosis and inflammatory response accompanies further decline in SI early after treatment with LVAD and HT; these levels improve long-term but do not supersede symptomatic HF. Gut microbial diversity was associated with measures of inflammation. Gut and oral microbial diversity and individual taxa were associated with SI.

Sarcopenia is highly prevalent among older adults and in HF patients, ranging from 20-50%, leading to reduced functional capacity and disability, increase in hospitalizations, health care costs and overall mortality^31^. The diagnosis of sarcopenia requires a comprehensive assessment of muscle mass, muscle strength, and physical performance. However, the complexity of sarcopenia assessment likely makes the formal screening for sarcopenia highly underutilized. The SI is a readily available biomarker of skeletal muscle mass that has been validated in several populations including critically ill^6^, older adults^32^, lung and kidney transplant recipients^7^, diabetic^33^ and cancer patients^34^. Moreover, SI was found to be a prognostic factor for short- and long-term survival and complications among cancer patients^35^.

Our study demonstrated a progressive decrease in SI with worsened HF class, representing skeletal muscle mass loss in the context of HF disease progression. The SI reached its nadir within the 1-month of treatment with LVAD or HT, likely reflecting the additional insult of surgery to the vulnerable advanced HF patient. A subsequent rebound in SI was observed 6-12 months following these interventions but only to the levels of symptomatic HF. These findings of worsened sarcopenia trends along with progression of HF and further decline following surgery are in line with prior studies^36, 37^. The incomplete resolution of skeletal muscle abnormalities following LVAD and HT has also been previously reported, utilizing skeletal muscle biopsies^38^ and more advanced imaging techniques^39^, and are concordant with clinical manifestations of residual diminished exercise capacity and cardiopulmonary reserve^40, 41^. The underlying mechanisms of muscle wasting in HF are complex and multifactorial. They relate to persistent levels of inflammation, oxidative stress, abnormal energy metabolism coupled with mitochondrial dysfunction, as well as hormonal changes, accompanied by transition of myofibers from type I to II and muscle atrophy^42^. During HF progression, altered gut microbial composition in the context of bowel wall congestion and hypoperfusion from HF, has been also hypothesized as a mechanistic link between HF, inflammation, and sarcopenia.

The observed relationship among HF, LVAD and HT patients with both inflammation and microbial diversity are consistent with our previous report and mirror patterns observed for SI, supporting the potential role of both inflammation and digestive tract microbiota in the development of sarcopenia. Specifically, there is a trend for decreasing microbial diversity (stronger for gut than oral) and increasing inflammation among more vs. less symptomatic HF and a notable drop in both gut and oral diversity, and increased inflammation in the ∼1-month following LVAD or HT. Consistent with rebounds observed for SI in months 3-12 following LVAD or HT, inflammation decreases and gut and oral microbiota increase. The altered gut diversity over time appeared to be driven by a depletion of *Firmicutes* and an enrichment of *Proteobacteria* immediately following LVAD and HT with a temporal shift back towards the composition observed in HF. While we have reported some of these patterns for gut microbiota and inflammatory biomarkers previously, this is the first report demonstrating consistent patterns for oral microbiota. The microbiome changes observed early after LVAD and HT while patients are still recovering from surgery in the hospital, are in line with prior reports of extreme gut and oral dysbiosis observed in the critically ill^43^.

Although our current findings are cross-sectional and cannot address temporality of relationships, it is plausible that sarcopenia is impacted by alterations in the gut microbiome. Recent animal models have shown the relationship between healthy gut microbiome and skeletal muscle (*gut-muscle axis*) and have identified SCFAs and inflammatory cytokines as responsible mediators^44, 45^. SCFAs are important gut microbiota byproducts that have been implicated in skeletal muscle mass health and function. SCFAs increase muscle glycogen levels, ATP production and promote gene expression involved in muscle protein synthesis through the insulin growth factor 1 pathway^46^. Decreased levels of SCFA were associated with chronic inflammation, likely further contributing to sarcopenia. In a large study of female twins, sarcopenia was inversely correlated with gut microbiota alpha-diversity and the relative abundance of *Faecalibacterium prausnitzii*, a well-known SCFA producer^47^. In the context of liver cirrhosis, Ponziani et al., reported that patients with cirrhosis and sarcopenia had depleted levels of *Methanobrevibacter, Prevotella, and Akkermansia* (generally considered as health-promoting taxa), and concurrent overgrowth of potentially pathogenic and inflammatory bacteria such as *Eggerthella* and *Klebsiella*^21^.

Our results support and extend prior literature. To our knowledge, the observation that *Roseburia inulinivorans* was associated with increased SI levels (after multivariable adjustment and adjustment for multiple comparisons) is the first report in HF, LVAD and HT patient. The finding is also consistent with its function as SCFA producer and several prior reports that this taxon is generally associated with healthier phenotypes^48^. Notably, despite prior studies suggesting a role for microbial AA producers in sarcopenia, none of our *a priori* identified taxa with known AA metabolism functionality were associated with SI. This could be a true null finding, but it could also reflect the limited ability of our data to detect specific AA producers as they might have low prevalence.

Our findings for salivary microbiota are highly novel as no prior study has broadly investigated the oral microbiota among HF patients or in relation to sarcopenia. Here we observe that findings for the oral microbiota are generally as strong as findings for gut microbiota. Patterns of salivary microbial diversity metrics were concordant with patterns observed for gut diversity (i.e., decreased diversity in more vs. less symptomatic HF patients and early after surgery, as well as positive correlations with SI). However, future studies are necessary to clarify the potential role of oral microbiota in sarcopenia as the taxa findings were somewhat inconsistent with current knowledge. For example, two *Prevotella spp*. were observed to be related to higher SI (less sarcopenia) levels despite the fact that many Prevotella spp. (including *Prevotella nanceiensis*) have been previously linked to oral inflammation and are enriched in diseased phenotypes^49^. In contrast, the presence of *Actinomyces naeslundii* was related to lower SI (more sarcopenia) values despite this taxon being regarded as a health-associated taxa in studies of both oral^50^ and systemic diseases^23, 51, 52^. These contradictory findings are notable given the large literature connecting oral microbiota to a variety of extra-oral cardiometabolic outcomes. Nevertheless, the broader literature on oral microbiota, systemic inflammation^53^, and cardiometabolic risk^54^ supports a potential role for oral microbiota in adverse HF outcomes, including sarcopenia. Notably, most prior literature connecting oral microbiota to extra-oral outcomes has focused on subgingival plaque microbiota whereas we currently have studied the saliva. Saliva microbiota composition is highly dynamic and likely reflects a mixture of various oral anatomical sites (supragingival, subgingival, tongue, cheek) all of which represent unique ecological niches with substantial diversity in microbiota composition^55^.

We observed that increased gut (but not oral) diversity is associated with lower levels of inflammation. We also observed that lower levels of inflammation are associated with increased SI (less sarcopenia). This pattern suggests that inflammation could be a mediatory linking gut diversity to SI although the cross-sectional nature of these data precludes formal mediation analysis and future longitudinal research is necessary to evaluate inflammatory mediation hypotheses.

Some limitations should be acknowledged. SI was used for the assessment of sarcopenia status. We did not perform formal assessments of muscle function or mass. However, we observed SI to be correlated with CT muscle cross-sectional area in a subgroup of our patients. We were unable to account for diet or other behavioral variables, and diet has been shown to be associated with microbiota measures and is plausibly linked to sarcopenia. Future studies that can account for these potential confounders will be important. The studied cohort was predominantly male, limiting generalizability. There is a clear difference in medication use among the groups. As expected, patients with HT adhere to an immunosuppression regimen, and we observed an uneven distribution of HF medications utilization among patients with Class IV HF, likely owing to reduced tolerance. Steroid use in HT can reduce systemic markers of inflammation and oxidative stress, as it has been shown in other disease states such as emphysema^56^.

In conclusion, we have observed declining SI trends with HF disease progression, the levels improve after treatment with LVAD or HT, but only to the levels of HF patients. Sarcopenia index covaries with inflammation and gut and oral microbiota in a similar fashion. Larger prospective studies (both observational and interventional) are warranted to address temporality of associations between skeletal muscle metabolism, inflammation, and digestive tract microbiome.

## Data Availability

All data is available upon request

